# Evolution of spray and aerosol from respiratory releases: theoretical estimates for insight on viral transmission

**DOI:** 10.1101/2020.07.23.20160648

**Authors:** P. M. de Oliveira, L. C. C. Mesquita, S. Gkantonas, A. Giusti, E. Mastorakos

**Affiliations:** Hopkinson Laboratory, Department of Engineering, University of Cambridge, Cambridge, UK; Mechanical Engineering Department, Imperial College London, UK

## Abstract

By modelling the evaporation and settling of droplets emitted during respiratory releases and using previous measurements of droplet size distributions and SARS-CoV-2 viral load, estimates of the evolution of the liquid mass and the number of viral copies suspended were performed as a function of time from the release. The settling times of a droplet cloud and its suspended viral dose are significantly affected by the droplet composition. The aerosol (defined as droplets smaller than 5 µm) resulting from 30 seconds of continued speech has o(1 h) settling time and a viable viral dose an order-of-magnitude higher than in a short cough. The time-of-flight to reach 2 m is only a few seconds resulting in a viral dose above the minimum required for infection, implying that physical distancing in the absence of ventilation is not sufficient to provide safety for long exposure times. The suspended aerosol emitted by continuous speaking for 1 hour in a poorly ventilated room gives 0.1–11% infection risk for initial viral loads of 10^8^-10^10^ copies/ml_l_, respectively, decreasing to 0.03–3% for 10 air changes per hour by ventilation. The present results provide quantitative estimates useful for the development of physical-distancing and ventilation controls.

## 1. Introduction

As the scientific debate on the droplet-related mechanisms allowing for the global spread of the SARS-CoV-2 virus evolves, attention has been shifting from *droplet* transmission towards *aerosol* as a possible additional route of transmission [1]. Illness caused by the inhalation of small-sized virus-laden droplets that can remain in the air long after being emitted by an infected (symptomatic or asymptomatic) individual, seems to explain a number of known outbreaks where safety measures put in place by local authorities were adopted, but contagion still occurred [2]. Due to the limited information on how respiratory droplet clouds evaporate while settling by gravity, the values and decay rates of the suspended viral dose emitted in respiratory releases are yet unknown. As a result, a great deal of uncertainty might be associated with current modelling practices used to define emergency control policies to minimise the damage caused by the COVID-19 disease as it develops. This work sheds light on the droplet-versus-aerosol debate by providing a theoretical characterisation of the time evolution of droplets from the moment of their emission, bridging these estimates to our current knowledge of the SARS-CoV-2 characteristics [3–7] in order to provide a metric of viral dose levels and decay.

Although significant efforts were made to investigate the time scales associated with single respiratory droplets undergoing the combined action of evaporation and gravity (starting from [8] and refined more recently by [9]), only recently a full experimental account of the actual range of droplet sizes produced in a respiratory release was possible. Due to fast atomisation processes happening at the mouth, vocal chords, and lungs, during breathing, speaking, or coughing, a wide range of droplets from sub-micrometer to millimetre size are emitted, requiring various experimental techniques combined to account for such wide-ranging sizes [10–12]. In addition to previous efforts, evidence that the evaporation process of a sputum droplet might be significantly affected by the droplet composition was shown very recently for a 10-µm diameter droplet [13]. As a multi-component droplet evaporates, the concentration of non-volatile components increases, affecting evaporation rates and ultimately dictating the final “equilibrium” diameter reached by the droplet.^1^ Hence, the final droplet diameters found in [13] were between 20–40% of the initial value, approximately, depending on the assumed concentration of protein modelled in the droplet. Such large variation leads to considerable errors in the modelling of *airborne* transmission, which assumes the resulting droplet nuclei to lie within the aerosol class, that is, below 5 µm [14].^2^ Additionally, such effects could also impact the short-range, short-time scale problem of *droplet* transmission.

Here, an evaluation of the evaporation and settling of droplet clouds emitted in respiratory releases is carried out, providing information on the time between the short-time scale problem of droplet transmission up to the long-time scale problem of aerosol transmission. State-of-the-art models are used for the prediction of evaporation and to account for droplet size distribution and concentration in two modes of droplet exhalation: by coughing and speaking. In this paper, we:

- Evaluate the size distribution and life-time of the suspended droplet cloud and the effects of ambient conditions on these quantities.
- Provide ensemble averages of mass and droplet number concentration in terms of their initial values at the emission source.
- Investigate, in the previously mentioned analyses, the evolution of different classes of droplets representing specific viral transmission scenarios: the whole of the suspended droplet cloud; only droplets within the aerosol category; and each of these two groups excluding “dry” droplet nuclei.
- Evaluate the emitted viral dose from sick individuals through coughing and speaking, and investigate the evolution of the suspended viable viral dose.

To demonstrate the impact of such metrics for physical distancing/ventilation, three canonical problems are considered: uniform flow from the emitter to the receptor, jet decay simulating the short-range flow pattern from the emission, and the well-mixed room focusing on the long timescales of the aerosol and on the risk of infection.

## 2. Methods

### (a) Droplet motion and evaporation

The sputum droplets are assumed to be spherical particles. The trajectory of the centre of mass is computed using the so called ‘Lagrangian’ approach where the equations of motion (balance of momentum) are solved for each particle independently. Both the gravity and buoyancy forces are considered together with the aerodynamic drag. All the other forces, such as Basset, Magnus and Saffman forces, as well as virtual mass are neglected [17]. The governing equations in a Lagrangian specification that describe the motion of the *k*-th droplet are as follows:

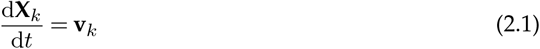

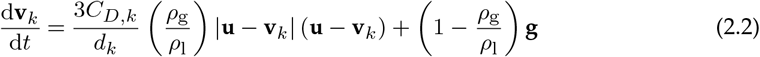

where *t* indicates the time, **g** is the acceleration of gravity, *d*_*k*_ is the diameter of the droplet, **X**_*k*_ and **v**_*k*_ are the position and velocity vectors of the droplet, **u** is the velocity vector of the gas, *ρ* is the density, and the subscripts g and l refer to the gas and liquid phase, respectively. The parameter *C*_*D,k*_ in the aerodynamic drag term of Eq. 2.2 is a function of the Reynolds number of the droplet Re_*k*_, calculated based on the relative velocity between the particle and the carrier phase, i.e., Re_*k*_ = *ρ*_g_*d*_*k*_ |**u** − **v**_*k*_| /*µ*_g_, where *µ*_g_ is the dynamic viscosity of the gas phase. Here, the Schiller-Naumann correlation for drag is used [18],

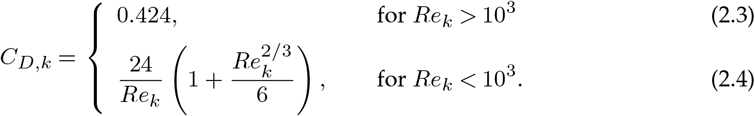

Further, mass and temperature equations for an evaporating droplet are given according to the model proposed by [19], assuming an infinite conductivity for the droplet. The evaporation model accounts for the effect of Stefan flow on heat and mass transfer between the droplet and the surrounding gas, assuming a dilute spray (i.e., volume fraction of the droplets below 0.1% – this allows us to use for each particle the models developed for isolated droplets) in an infinitely large domain:

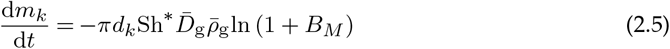

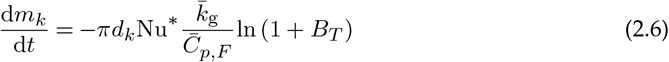

where 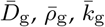, and 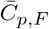 are the average binary diffusion coefficient of the mixture, density, thermal conductivity, and specific heat in the film. The parameters *B*_*M*_ and *B*_*T*_ are the Spalding mass and heat transfer numbers, and the modified Nusselt and Sherwood numbers are given as,

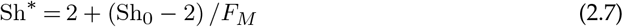

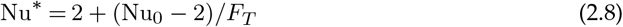

These are calculated based on the actual Nusselt and Sherwood numbers, obtained with the widely used Frossling’s correlations [20], and then corrected for the film thickness of the surrounding gas by the correction factors *F*_*M*_ and *F*_*T*_ proposed in [19]. Given the low Biot number of the droplets in all conditions, Bi_*k*_ = *hd*_*k*_/(2*k*_g_) < 0.1, where the convective heat transfer coefficient *h* is computed from the Nusselt number, a convection-controlled process is assumed and, in turn, a uniform internal droplet temperature *T*_*k*_ can also be assumed. The rate d*T*_*k*_/d*t* can be obtained by an energy balance at the droplet,

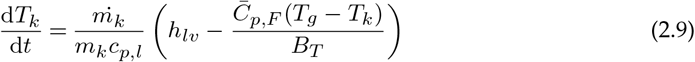

In the present calculations, it is assumed that the air is initially at a homogeneous temperature *T*_*a*_ and pressure *p*_*a*_, and contains moisture characterised by the volume fraction of water vapour *x*_*w*_, which is related to the relative humidity by *x*_*w*_ = RH *p*_*w,sat*_/*p*_*a*_. Such conditions constitute the far-field (i.e., a region sufficiently far from the droplet surface, where the evaporation process has negligible effect) boundary conditions for droplet computations. The initial conditions of the spray are described next, in Sec. 2(c).

In an initial approach, the liquid composition is considered to be pure water. To mimic the formation of a “droplet nucleus” (i.e. what remains after evaporation has ended), the effect of non-volatile content diluted in the droplet is implemented by simply limiting evaporation of each droplet down to 6% of its initial mass, that is, *m*_*k*_/*m*_*k*,0_ = 0.06. This value is representative of the equilibrium diameter for the range of relative humidity between 20% and 80% verified by [16] for a model sputum droplet containing 9 mg/ml of NaCl, 0.5 mg/ml surfactants, and high protein concentration of 76 mg/ml. Nevertheless, it is worth noting that the equilibrium diameter as evaluated in the model is approximately 50% smaller for a model sputum droplet with low protein concentration (3 mg/ml) within the same relative humidity range [16]. In Sec. 2(b), we provide the additional models to evaluate the effect of the droplet composition on evaporation. It should be noted, however, that neither approach considers the effect of hygroscopic growth of the droplet nuclei that may happen at high relative humidity for a recently emitted droplet, or could also be observed in a droplet nucleus undergoing changes in the relative humidity in the environment.

### (b) Other effects on evaporation

It has been mentioned that the effects of solute and of the droplet curvature can determine the equilibrium size of the aerosol [13]. To account for the increase in the water vapour pressure due to the curvature of the droplet surface, Kelvin’s equation [21] is employed,

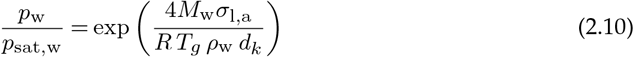

where *M* is the molecular weight, *σ* is the surface tension, *R* is the universal gas constant, and the subindex *w* represents water. The effect of the surface curvature may be enhanced due to a surface tension increase in the presence of NaCl and other components in the liquid [22], and at the same time be suppressed by the presence of surfactants which lowers the surface tension [23]. Thus, the surface tension of pure water was used as an approximation.

The saturation pressure of the water at the droplet interface is decreased by the presence of soluble as well as by insoluble substances in the droplet. To account for this in a multicomponent sputum droplet, Eq. 2.10 becomes, [21,24,25],

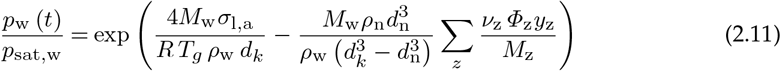

where *ν*_z_ is the number of ions into which a solute molecule *z* dissociates, *ϕ*_z_ is its practical osmotic coefficient, and *y*_z_ its mass fraction in terms of the total dry mass. We consider the presence of NaCl, proteins, and surfactants in the liquid. Details for the calculation of the ion-interaction parameterisation of *ϕ*_z_ for NaCl and osmotic pressure parameterisation of *ϕ*_z_ for BSA proteins are given in [24]. The latter was also used for the evaluation of that parameter in dipalmitoylphosphatidylcholine (DPCC) surfactant, considered here.

Finally, assuming the water as the only volatile component in the liquid droplet, it is possible to demonstrate that the rate of change of the mass fraction of water in the droplet due to evaporation is,

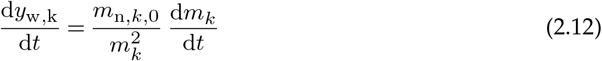

where *m*_n,*k*,0_ is the initial mass of non-volatile (dry) components in the droplet *k*, respectively.

Three different initial liquid compositions were considered, reflecting directly on the evaluation of the water vapour pressure at the droplet surface (Eq. 2.11): pure water, a high protein sputum and low protein sputum. The initial concentrations and molecular weight of each component are given in Table (b). The sputum compositions used in this study are those considered in [13], which are low protein content found in the nasal surface airway fluid [26], and high protein content found in breath aerosols [27], both considering a small concentration of DPPC as surfactant [28]. A homogeneous composition across all droplet sizes is considered for each case, although in reality different droplet compositions may be found depending on the origin of the droplets.

### (c) Droplet size distribution and viral activity

Size distribution of droplets escaping the mouth, associated with coughing and normal paced speaking, are given as probability density functions (pdfs) of droplet diameter. These functions were obtained using the tri-modal log-normal distribution provided by [12]. The model was named by the authors as the bronchiolar-laryngeal-oral tri-modal model, as it considers droplet production associated with three distinct modes: one occurring in the lower respiratory tract, another in the larynx, and a third in the upper respiratory tract and oral cavity, respectively. Here, we consider an initial droplet distribution in the range 1 µm–1mm. The number concentration of droplets of size *k* produced in each of these modes is given as a sum over each mode *i* [12]:

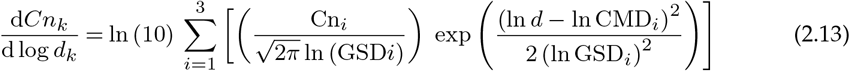

where the droplet number concentration Cn_*i*_, geometric standard deviation GSD_*i*_, and count median diameter CMD_*i*_ for *i*-th mode are given in Table 2. The total number and liquid concentrations per unit volume of exhaled gas can be directly obtained by sum of Cn_*i*_ and Cm_*i*_, respectively, for the three modes. The mass concentrations were evaluated by [12] assuming perfectly spherical droplets and the density of pure water.

**Table 1.** Non-volatile composition of pure-water, high-protein sputum, low-protein sputum droplets, and molecular weight and density of each component.

| | $c$ (mg/ml) | | | $M_w$ | $\rho$ |
| --- | --- | --- | --- | --- | --- |
|  | pure water | h-p sputum | l-p sputum | (g/mol) | (kg/m<sup>3</sup>) |
| NaCl salt | 0 | 9 | 9 | 58.4 | 21600 |
| BSA protein | 0 | 76 | 3 | 66500 | 1362 |
| DPPC surfactant | 0 | 0.5 | 0.5 | 734 | 1000 |

**Table 2.** BLO model parameters from [12].

| i | 1<br>(B mode) | 2<br>(L mode) | 3<br>(O mode) |
| --- | --- | --- | --- |
| Speaking |  |  |  |
| $Cn_i$ (#/cm<sup>3</sup>) | 0.069 | 0.085 | 0.001 |
| $CMD_i$ (μm) | 1.6 | 2.5 | 145 |
| $GSD_i$ (-) | 1.3 | 1.66 | 1.795 |
| $Cm_i$ (μg/cm<sup>3</sup>) | 0.21 | 2.2 | 7500 |
| Coughing |  |  |  |
| $Cn_i$ (#/cm<sup>3</sup>) | 0.087 | 0.12 | 0.016 |
| $CMD_i$ (μm) | 1.6 | 1.7 | 123 |
| $GSD_i$ (-) | 1.25 | 1.68 | 1.837 |
| $Cm_i$ (μg/cm<sup>3</sup>) | 0.22 | 1.09 | 69000 |

We model the virus decay in a droplet assuming an exponential decay at a rate *λ* such that [29],

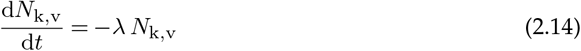

where *N*_k,v_ is the total number of *viable* viral copies (i.e., plaque forming units, or PFU) in a single droplet *k*. As an approximation, an exponential decay constant of *λ* =0.636 h^−1^ is used for all droplet sizes and compositions. This value was obtained from the experiments performed by [29], which verified the stability of the SARS-CoV-2 virus as an aerosol, formed of 5 µm droplets released in 65% relative-humidity ambient air. In that work, a virus half life of 1.09 h was observed. One should note that with this approach, local droplet effects such as change in solute concentrations with evaporation on virus activity are neglected. Further, the initial viral copies in each droplet is defined as,

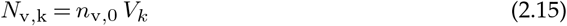

where *V*_*k*_ is the volume of the droplet. The initial viral load *n*_v,0_ has been recently reported for SARS-CoV-2, with typical values found in sputum and throat swabs ranging between 10^9^ to 10^4^ copies/ml_l_ as the disease develops in sick individuals [4,6,30], who were tested daily from the on-set of the symptoms of the disease and up to several days later. Still, values as high as 10^10^ copies/ml_l_ were observed [30], and even 10^11^ copies/ml_l_ in patients who recently died [6].

Additionally, evidence of transmission caused by an asymptomatic individual has also been suggested; following the outbreak, a viral load of 10^8^ copies/ml_l_ was detected in the individual’s sputum [5].

The total suspended viable viral copies and viable viral load in copies, or plaque forming units, per unit volume of liquid are:

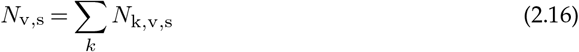

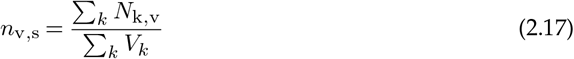

Finally, in order to put the results of Sections 3(a)–(b) in context of physical distancing rules, the infection dose of virus in viable copies needed per individual 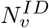 leading to the disease must be defined as a reference. This value is normally reported as the amount of plaque forming units which will result in a given percent responses (illness) over X% of a population – e.g. 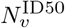 for infection of 50% of a population. In the lack of these values for SARS-CoV-2, we used the infection dose model by [31] for SARS-CoV-1 due to the claimed similarity of this virus with SARS-CoV-2 and as it is in the same genetic group as other coronaviruses such as HCoV-229E (human common cold), MHV-S and HEV-67N (animal coronaviruses) for which the model also demonstrated a good agreement with experimental data [31]. The intervals of 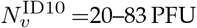 PFU and 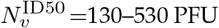 PFU are reported for a 95% confidence interval.

## 3 Results

### (a) Ensemble averages and time scales of suspended droplet clouds

The settling of human exhaled droplet clouds is analysed in the absence of turbulence and of mean air flow, so that the effects of gravity and evaporation on various ensemble quantities of interest can be independently investigated. The Lagrangian framework, given in Sec. 2(a), is considered in one (vertical) dimension and droplet clouds for two exhalation modes, speaking and coughing, are released at the height of the emitter’s mouth (1.5 m) and then let settle by gravity while evaporating in ambient air. In this section, the effect of composition was not taken into account, hence droplets were considered as pure water with evaporation limited down to the equilibrium size corresponding to 6% of its initial mass.

So as to provide an overview of the evaporation-settling process, the probability density functions (pdfs) of droplet size for coughing and speaking are provided in terms of the time after emission for an ambient condition of *T*_a_=20°C and RH=80% (Fig. 2). In this analysis, all droplets or droplet nuclei that remain suspended in air in a given time are considered. This ensemble average is denoted by the subscript s in the quantities of interest (e.g. *Ψ*_s_). The tri-modal bronchiolar-laryngeal-oral characteristic pdfs of droplets produced during speech and cough [12] are given at *t* = 0 s. Within one minute, most of the large droplet portion of the distribution (100 µm–1 mm) progressively disappears as these droplets reach the ground. One hour after emission, only droplets smaller than 5 µm remain suspended, that is, those typically defined as aerosol. Interestingly, at this point, the total number of particles in the sub-micrometre range is an order of magnitude higher for coughing than speaking.

**Figure 1.**
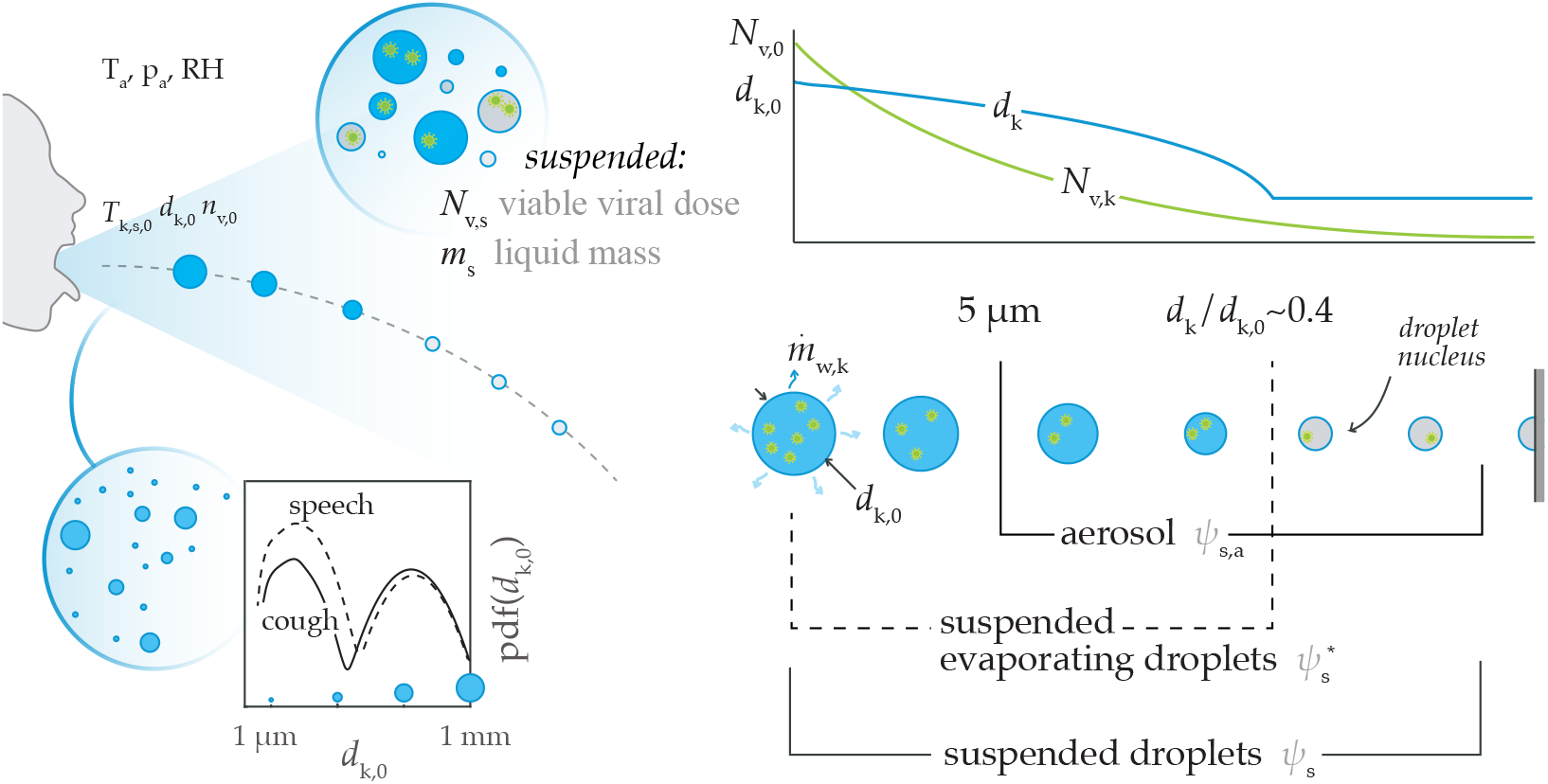
Illustration of the problem and main parameters.

**Figure 2.**
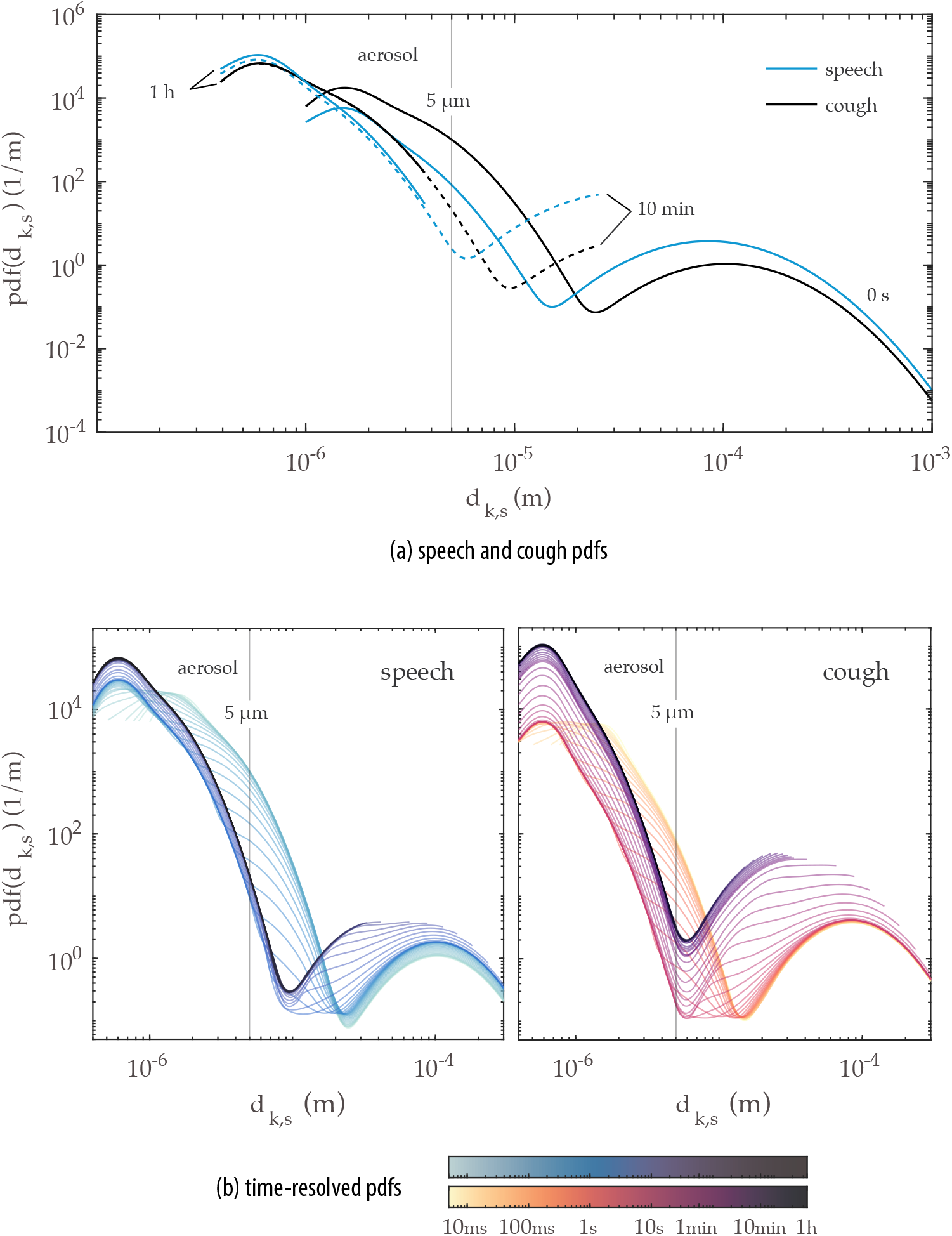
Evolution of the probability density function of droplet size from speaking and coughing, where (a) shows 0 s, 10 min, and 1 h pdfs, and (b) shows detailed time-resolved pdfs betwenn 0–1 h coloured in a log scale – T=20°C, RH=80%.

Next, we evaluate the pdfs of droplet size for a suspended *and evaporating* droplet cloud – that is, considering only droplets that have not yet reached their equilibrium size or, in other words, ignoring any droplet nuclei. This condition is defined by the superscript ^∗^, as 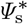. Figure 3 shows the evolution of speech-emitted droplet pdfs evaluated for (a) the total suspended droplet cloud and (b) those evaluated for the suspended cloud minus the droplet nuclei. These are reported for the same ambient condition of Fig. 2. A fast change to the coughing and speaking droplet clouds occurs within the first second from emission, as droplets of the order of 1 µm fully evaporate. These small droplets quickly reach their minimum (equilibrium) size, hence determining early-on in the process the aerosol dispersion that might remain suspended in the air for much longer times. The effect of evaporation on droplet size and the resulting time scales for full evaporation for each droplet class can be more easily seen in 3(b), as the lower limit of the given pdfs moves towards higher values as evaporating droplets reach equilibrium. The droplets within the bronchiolar mode of the probability density curve, centred at approximately 1-2 µm, have a 10-ms evaporation time scale, while those in the laryngeal mode a characterised by a longer, 1-s time scale. Moreover, Fig. 3b illustrates that the long-time scale problem may be reduced to 10-s time scale and droplets produced in the oral-mode in a virus transmission problem through droplets produced by normal, paced speech and where the presence of water is vital to the viability of the virus in the droplet. This could be, for example, due to an increasing concentration of surfactants, salts, and other non-volatile components that can enhance viral decay [16]. In that case, droplet sizes range between 10–100 µm, as no droplets within the aerosol class are observed after 100 ms and any droplets larger than 100 µm are quickly deposited by gravity.

**Figure 3.**
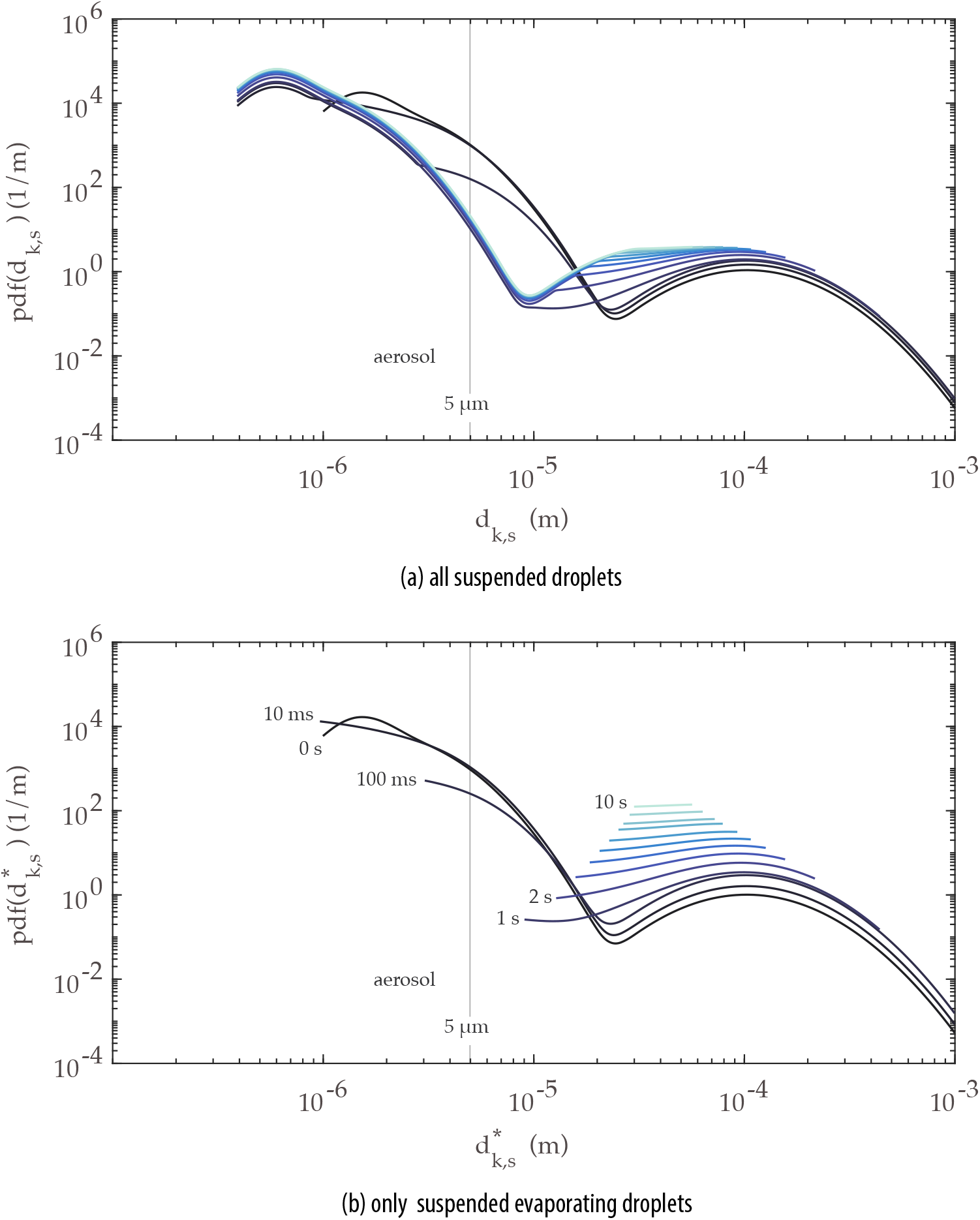
Evolution of the probability density function of droplet size from speaking for various times after droplets are exhaled. In (a) all suspended droplets are considered, and (b) only droplets that have not yet reached equilibrium – T=20°C, RH=80%.

Figures 5 to 7 provide insight on the time scales needed for viral removal from the ambient air due to settling of the droplets through gravity. First, the effect of relative humidity is shown on the evolution of two ensemble quantities of the droplet clouds exhaled by speaking and coughing: mass and number of suspended droplets, both quantities normalised by their respective values at *t* = 0 (Fig. 5). For reference, the total mass of liquid emitted in a single cough, lasting approximately 0.5 seconds, is equal to that of a 30-s speech; as evaluated from the concentration distributions provided in [12] and the exhalation gas flow rates for speaking (0.211 l/s) [32] and coughing (1.25 l/s) in [33]. For both exhalation modes, a significant portion of the emitted droplet cloud mass lies in the large droplets. In fact, 99% of the initial mass is concentrated in the droplet diameter range between 100 nm–1 mm (Fig. 4). Thus, it is not surprising that the decay of the total mass suspended relative to the initial emitted liquid mass (*m*_s,a_/*m*_0_, Fig. 5) behaves similarly in both exhalation modes. Nevertheless, what is interesting to observe in terms of the effect of evaporation and relative humidity on *m*_s,a_/*m*_0_ concerns the ensemble quantities for the resulting aerosol. As discussed previously, evaporation affects the small droplets in the aerosol within the first second after emission. Thus, the effect of lower (far-field) relative humidity which enhances evaporation is only noticed in the aerosol droplets within that time frame. While a decrease of the relative mass occurs due to the evaporation of the droplets in the aerosol class, a combined increase in the aerosol mass and droplet number also occurs as larger droplets evaporate and shrink below 5 µm. Further, the number droplets within the aerosol class relative to the initial number of droplets rises from 95% to 99% due to this effect, while for coughing only a small increase is observed (from 91% to 92%, roughly). The increase in *N*_s,a_/*N*_0_ due to evaporation in speech-emitted droplets is typically neglected but can be relevant when one assumes aerosol transmission only, say, in a well-mixed room type of ventilation problem (similar to [2]) where the intention is to evaluate risk of infection. This is aggravated by the fact that, relative to the initial emitted cloud, the number of droplets and the respective suspended mass after one hour is an order of magnitude higher for speaking than for coughing. This analysis illustrates the risk associated with constant speaking in closed environments (as for example, in a lecture hall) due to a higher mass fraction of liquid resulting in small droplets during speaking (Fig. 4).

**Figure 4.**
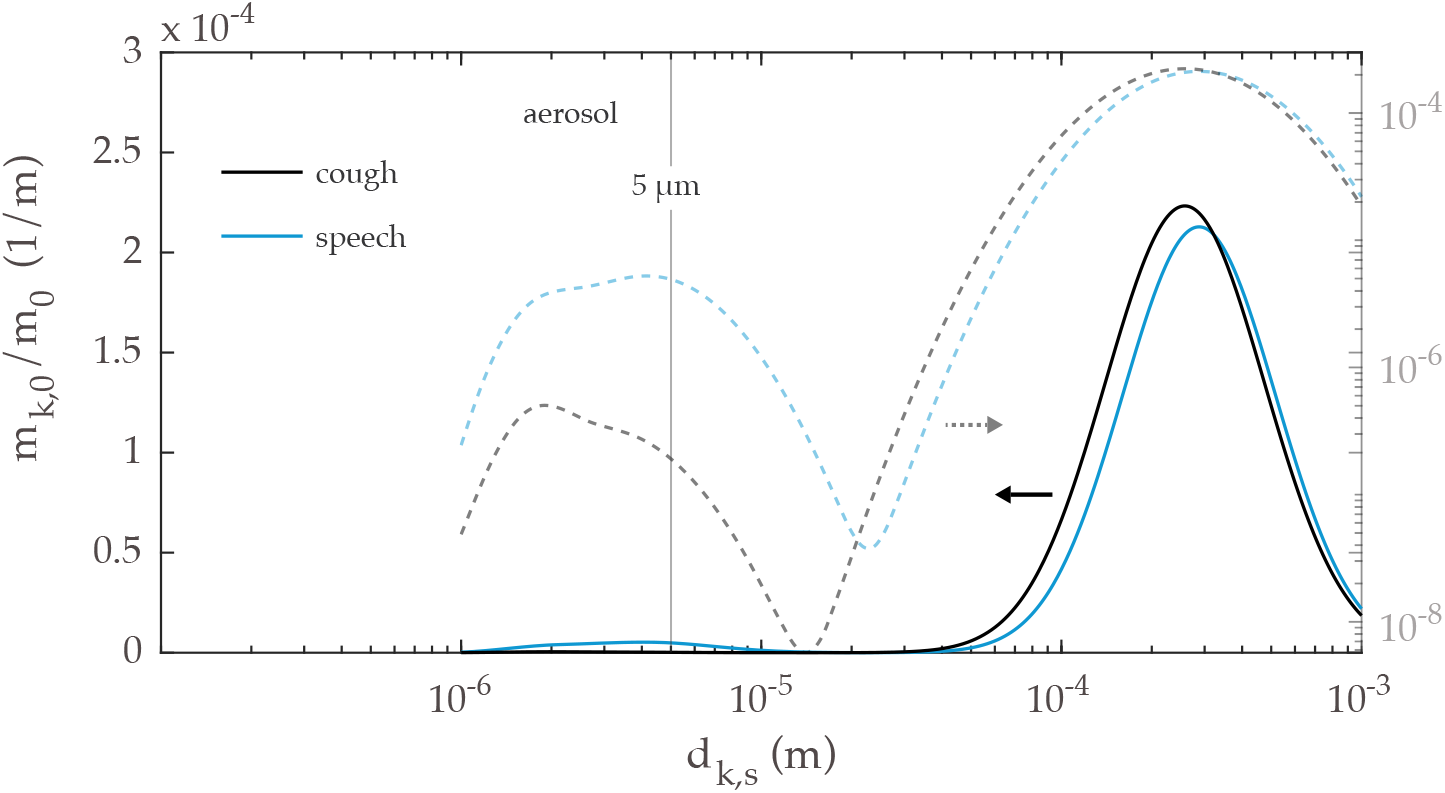
Initial mass in each droplet size category normalised by the total emitted mass of sputum in coughing and speaking. Dark lines represent the curves plotted in a linear vertical scale (left side y-axis), while the lighter colours represent the curves plotted in a vertical log scale (right side y-axis).

**Figure 5.**
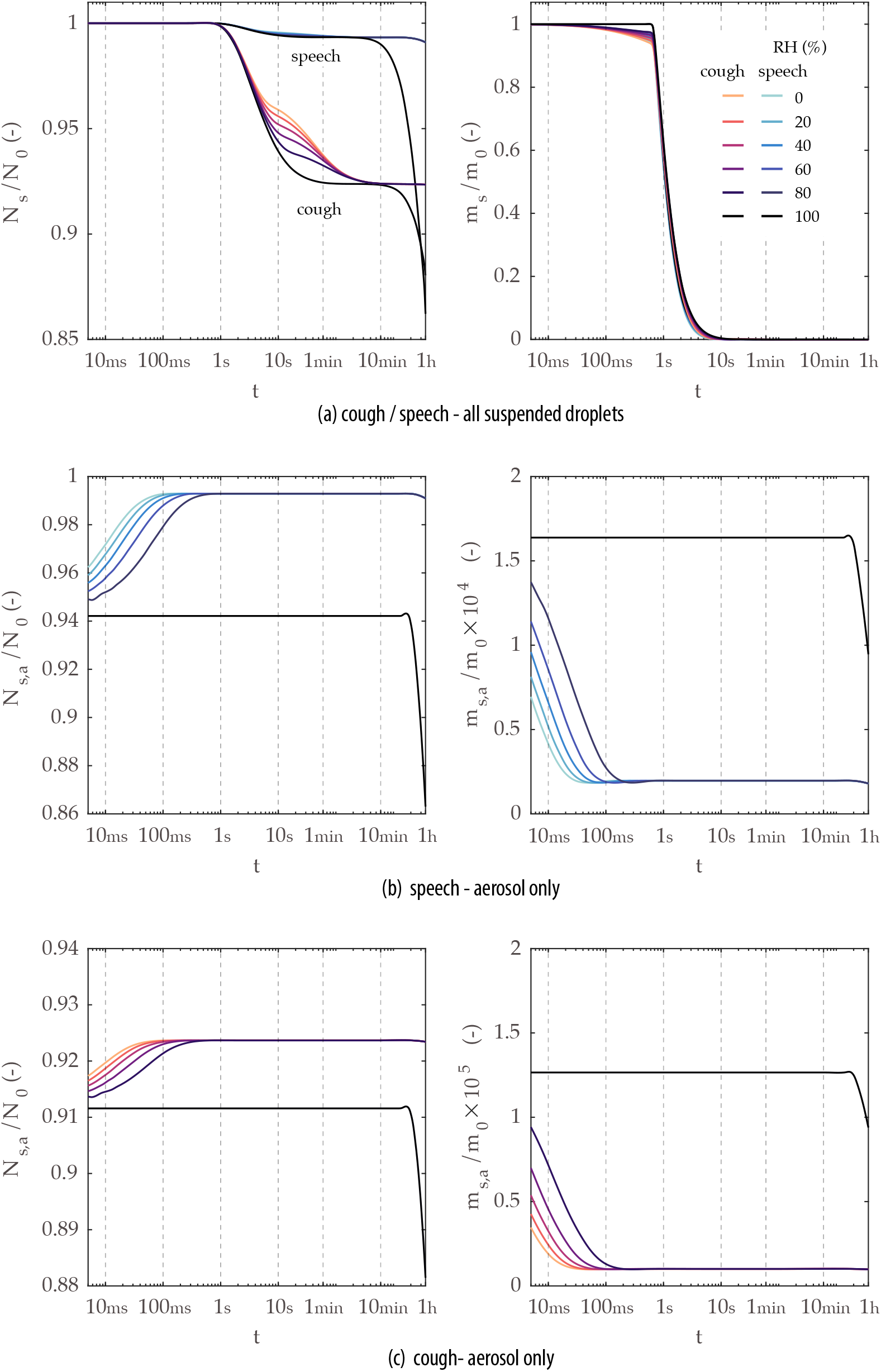
Time evolution of ensemble quantities (total mass and number) normalised by the respective value at *t* = 0 for sprays exhaled by speech and cough – T=20°C, RH=0–100%. In the top row, ensemble quantities shown are evaluated for all suspended droplets, while in the middle and lower low the quantities are calculated considering only droplets below 5 µm. More detail on the time evolution of *m*_s_/*m*_0_ is provided in Fig. 6.

The effects of evaporation and gravity on the time evolution of the ensemble quantities of interest are now discussed with a focus on transmission through evaporating droplets only. In Fig. 7, the relative mass and droplet number are evaluated for the total droplet cloud minus droplet nuclei, as well as for droplets in the aerosol minus droplet nuclei. In contrast with Fig. 5, where droplets and droplet nuclei are considered, full decay of the evaporating aerosol mass occurs roughly between 100–500 ms following exhalation. In terms of total mass emission, virtually no change is observed relative to the value shown in Fig. 5, as the mass decay is mostly controlled by deposition of large droplets on the ground, as previously discussed.

**Figure 6.**
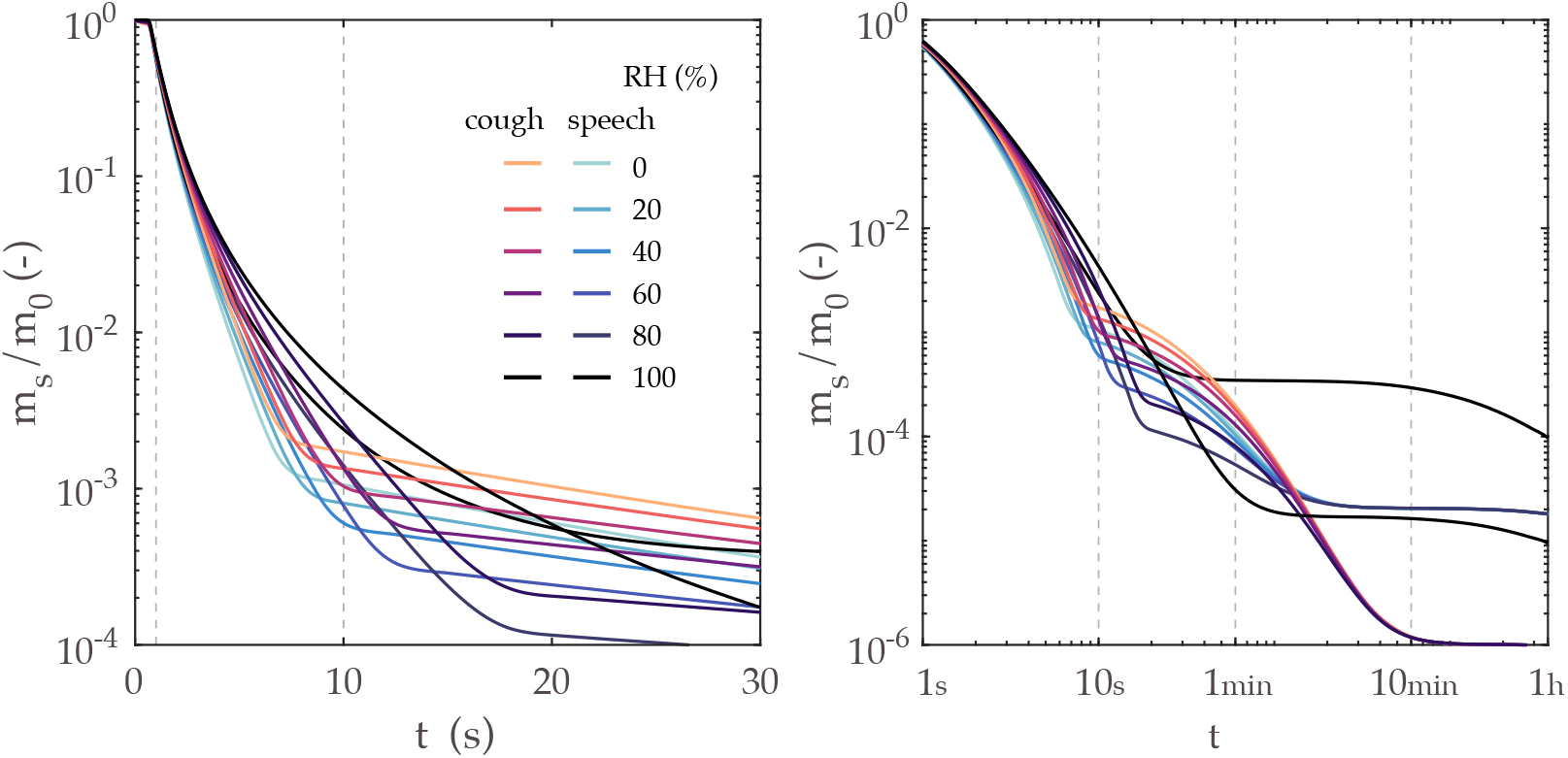
Detail of the time evolution of total mass normalised by the respective value at *t* = 0 shown in Fig. 5. The left-hand side plot is given in a linear time scale up to 30 s, while the right-hand plot is given for a log scale up to 1 h.

**Figure 7.**
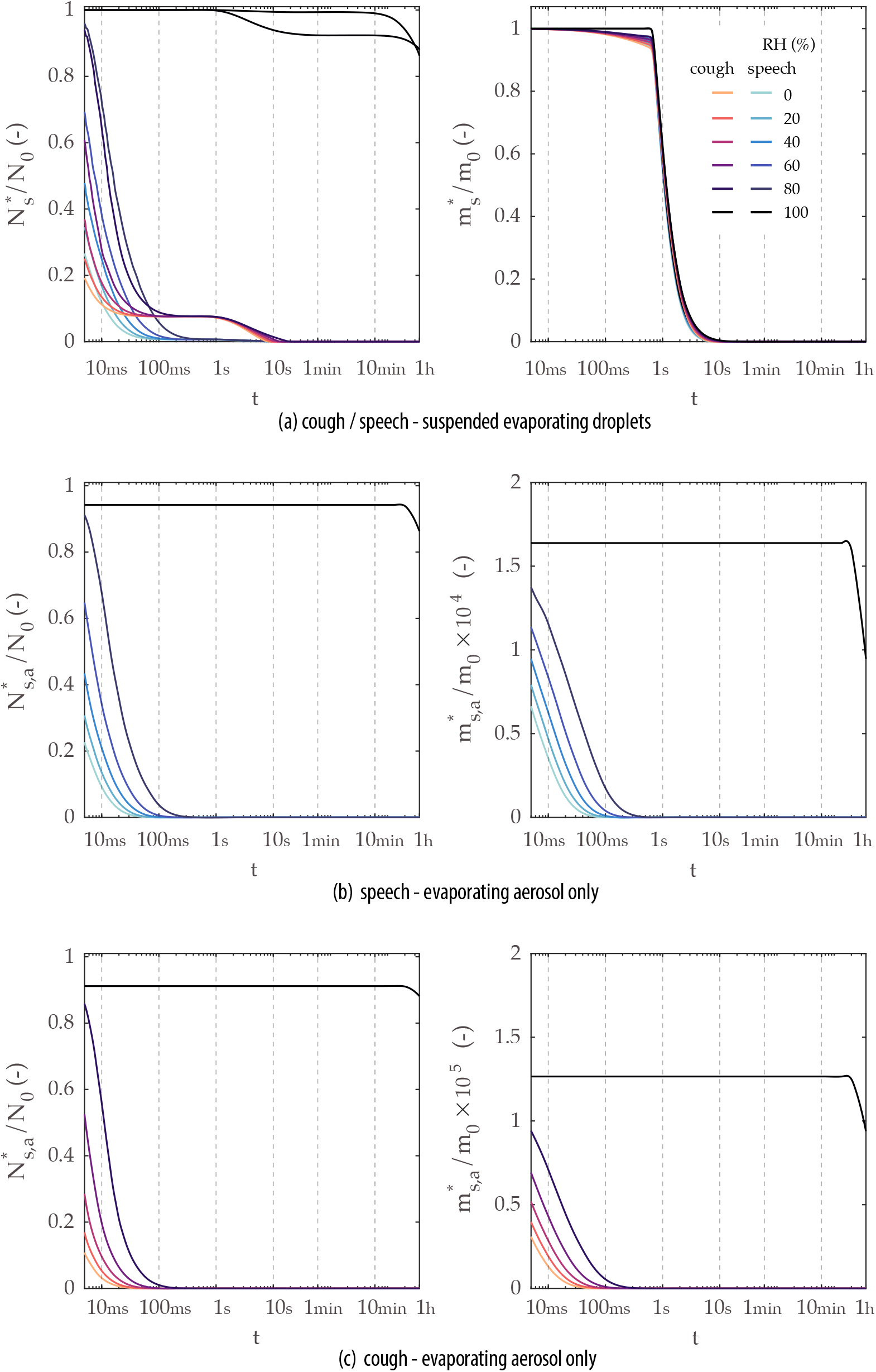
Time evolution of ensemble quantities (total mass and number) normalised by the respective value at *t* = 0 for sprays exhaled by speech and cough – T=20°C, RH=0–100%. The quantities only consider evaporating droplets, that is, droplets that have not yet achieved equilibrium with ambient air. In the lower row, ensemble quantities shown are calculated considering only droplets below 5 µm.

### (b) Considerations on the effect of droplet composition

In an attempt to provide an assessment of the uncertainty associated with the results given in Sec. 3(a), we assess the effect of different droplet compositions on evaporation and, in turn, on the resulting equilibrium size of the droplets. This is discussed in terms of the ensemble quantities and time scales discussed previously. First, this issue is analysed in the context of a single droplet. Figure 8 illustrates the problem through (a) evaporation and (b) settling of a 10-µm diameter droplet. The colour lines represent different relative humidities (RH=0–100%), while the short dash, long dash, and continuum lines represent each of the compositions used: that of pure water, high-protein and low-protein sputum, respectively. For the sake of comparing the sputum droplets with pure-water droplets in Fig. 8, no hard limit is imposed on the minimum droplet size of pure water in this case. Hence, water droplets undergo full evaporation in less than a second, except in the case of 100% relative humidity. As the droplet evaporates, the concentration of non-volatile components increases in the droplet, thus reducing the the vapour pressure at the droplet interface. Thus, a larger equilibrium diameter is reached for the case of high-protein sputum composition. As expected, evaporation is limited by the vapour pressure in the ambient. Thus, the closer to saturated air (RH=100%), the higher is the equilibrium diameter of the droplet. Although a large variation of the equilibrium diameter was expected, observed here between 20% and 50% of the initial diameter, its impact on the settling time is quite significant. This can be seen for the 10-µm droplet by taking the respective time at the crossing of each line with the x-axis in Fig. 8b.

**Figure 8.**
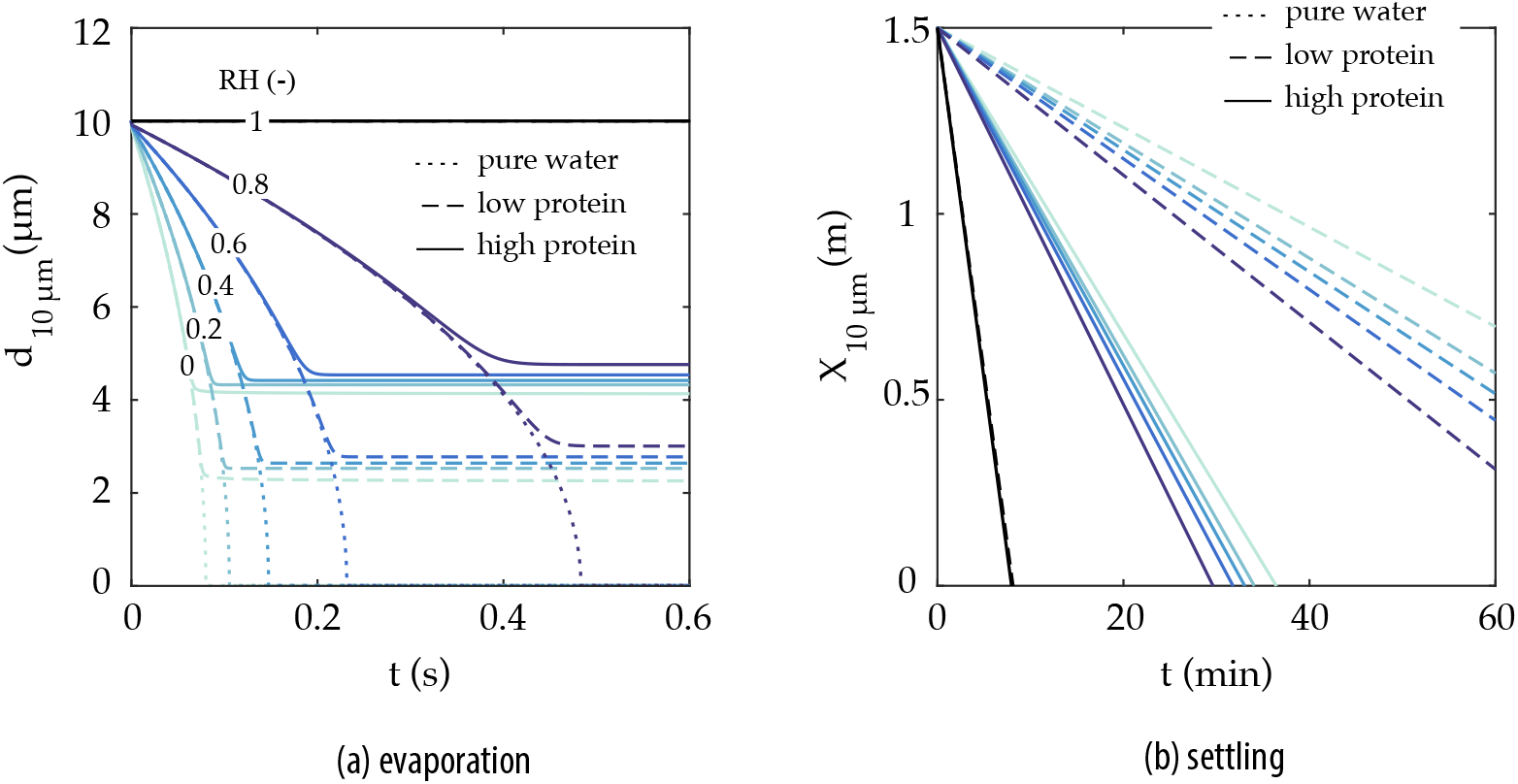
Impact of the droplet composition modelling approach (pure water; sputum 9 mg/ml NaCl, 3 mg/ml protein; sputum 9 mg/ml NaCl, 76 mg/ml protein) on the (a) evaporation and (b) settling of 10 µm droplets under different relative humidities. The relative humidity is shown in colour (RH=0–100%) – T=20°C.

As discussed in the context of a single 10-µm droplet in Fig. 8a, the combined effect of composition and relative humidity can have a major impact on the settling time of a droplet. Next, Fig. 9, we discuss this issue for the entire range of droplet sizes typically found in respiratory releases. The absolute settling time is shown in (a) in terms of the droplet size and for high- and low-protein sputum droplets. The relative difference between the two is given on the right-hand axis. Settling times of droplets in the 1–50-µm range are strongly affected by composition, while this effect is negligible in droplets larger than 100 µm. In the transition range between 50–100-µm, the deposition and evaporation time scales are very similar, hence the effect of composition swiftly disappears once the settling time due to gravity becomes much shorter than the droplet evaporation time scale. Further, Fig. 9b shows the aerosol droplet size distribution for coughing and speaking, where the total number of droplets for each size normalised by the total number of emitted droplets is given. By lowering the equilibrium diameter, more droplets are found in the aerosol category. The shape of the distribution is similar between the two droplet compositions, with the high-protein sputum droplets just shifted towards smaller diameters.

**Figure 9.**
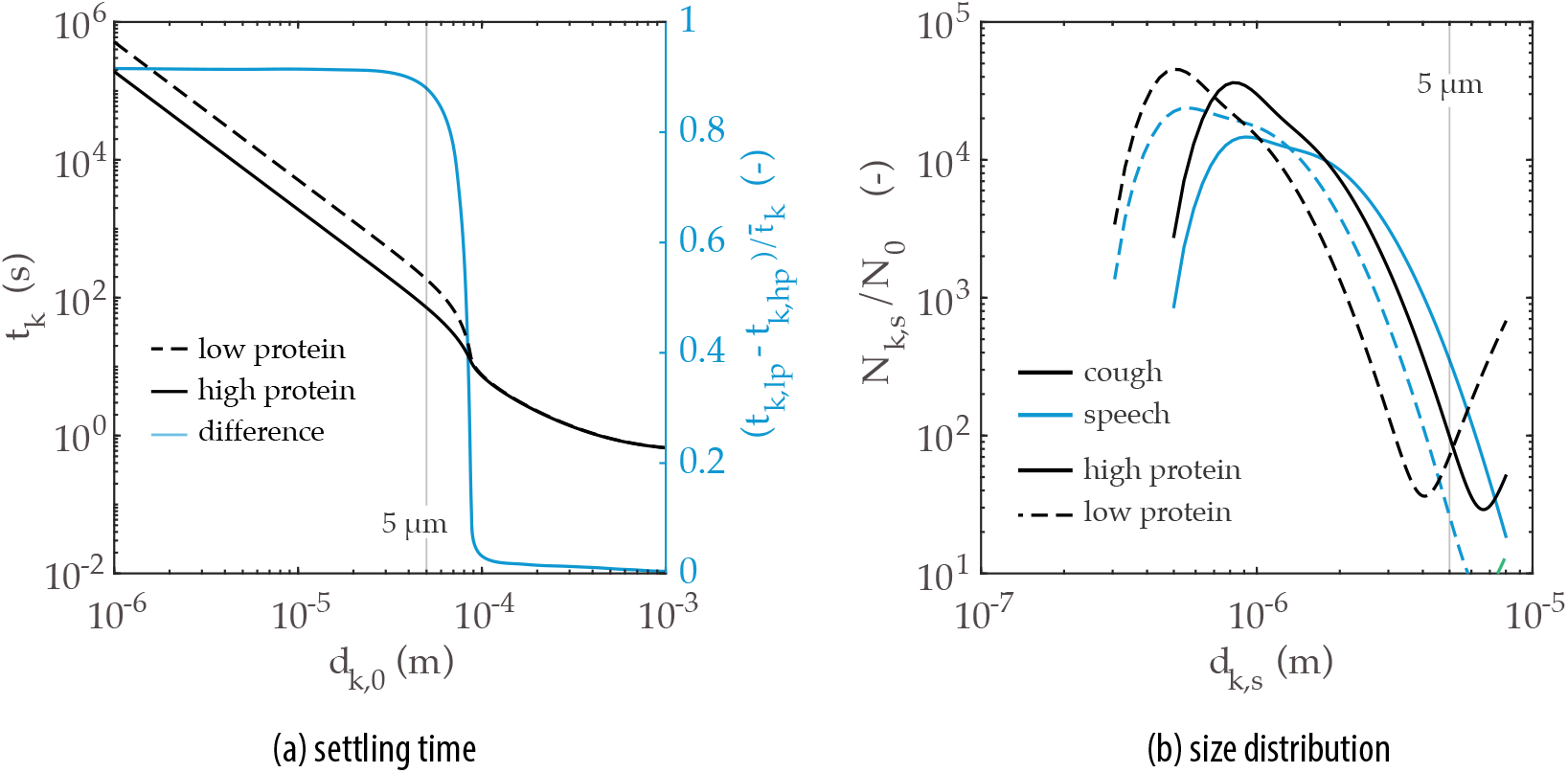
(a) Settling time (left axis) and its respective relative difference (right axis) of droplets and (b) probability density functions of droplet size normalised by the initial number of droplets at *t* = 10 min for coughing and speaking, both presented in terms of the saliva composition: low protein (sputum 9 mg/ml NaCl, 3 mg/ml protein) and high protein (sputum 9 mg/ml NaCl, 76 mg/ml protein) – T=20°C, RH=80%.

There are key implications of such findings. Although, at a first glance, one would be tempted to neglect the composition effect in the gravity-controlled short-time scale viral transmission problem (i.e. the ballistic and exhaled gas-driven motion of large droplets), significant differences in settling time and size in the 50–100-µm range can be critical. Such droplets might remain longer in air at face height, being more likely to be inhaled by someone near a sick individual. This problem has a direct impact on the uncertainty of methods to evaluate the safe physical distancing and, therefore, should be further investigated for better understanding, for example by means of high-fidelity flow simulations. Besides the short-time scale implications, the large differences in deposition times of droplets in the aerosol class, as an effect of both relative humidity (Fig.8a) as well as composition (Fig. 9b), might also affect the long-time scale problem, that is, the virus removal by ventilation in indoor spaces. This large difference can be clearly seen at large times in terms of *m*_s_/*m*_0_ and *N*_s_/*N*_0_ (Fig. 10) not only due to the significantly longer deposition times of such droplets, but also due to the difference in the mass that can remain suspended. Some of these issues are explored next, in Sec. 3(c)

**Figure 10.**
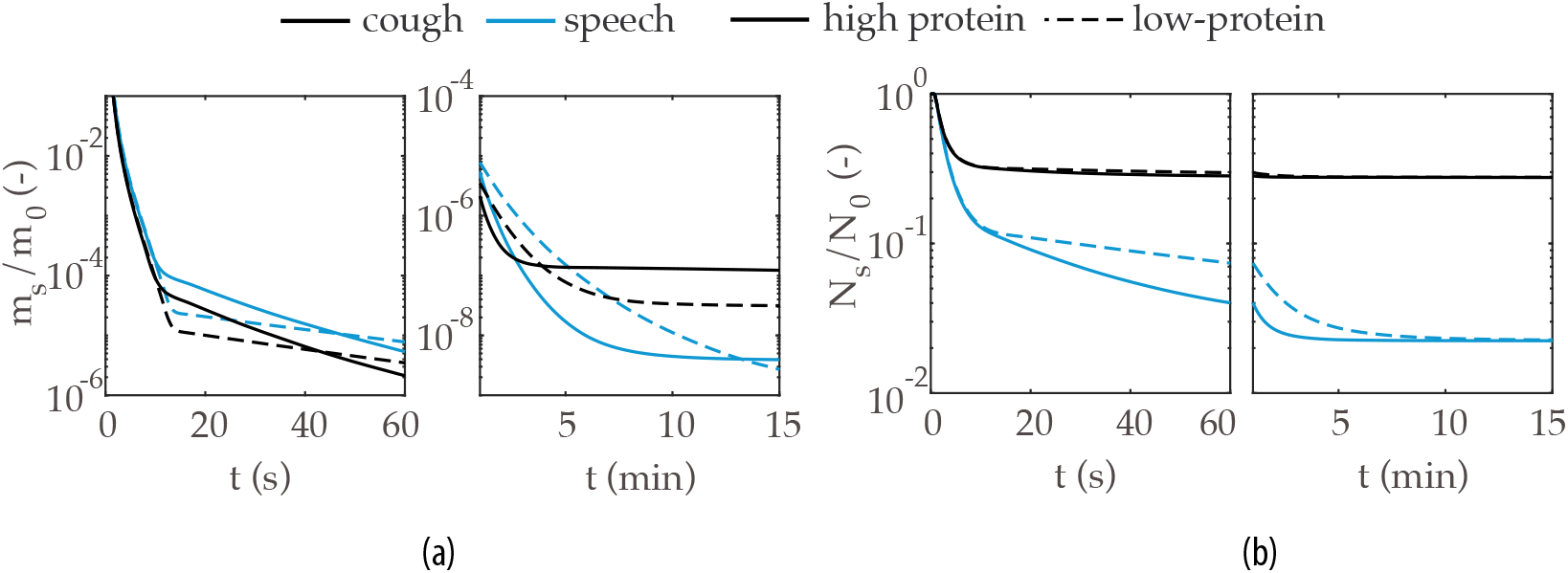
Time evolution of (a) total mass and (b) total droplet number, all normalised by their respective values at *t* = 0, in terms of the composition: low protein (sputum 9 mg/ml NaCl, 3 mg/ml protein) and high protein (sputum 9 mg/ml NaCl, 76 mg/ml protein) – T=20°C, RH=80%.

### (c) Suspended viable virus

In this section, the evolution of the total number of suspended viable viral copies is evaluated for speaking and coughing. We consider a single cough in which all droplets are assumed to be released instantly, and continuous speaking in which the droplets are considered as uniformly emitted over its duration. Assuming a short 0.5-s cough with a mean volumetric gas flow rate of 1.25 l/s [33], the total emitted liquid can be directly obtained from the total exhaled volume of gas and the liquid concentration values available in Table 2. Similarly, we consider the case of continuous, paced speech at an average volumetric gas flow rate of 0.211 l/s [32]. Hence, both exhalation modes result in the *same* emitted liquid mass. Additionally, for this analysis, we consider two sputum compositions (high-and low-protein sputum) and assume a range of low and high viral loads at the emission source to be that of a symptomatic person in: (i) a few days after the symptoms appear (typically 10^8^ copies/ml_l_) and (ii) at a severe stage in the disease (10^11^ copies/ml_l_). The evolution of the total number of suspended viable copies of virus *N*_s,v_ is given for those conditions and 40 and 80% relative humidity (Fig. 11). As a reference value, the infection doses 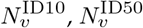, and 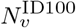 are also given. For speaking, an additional plot is given with a time scale relative to the end of the 30-s speech (Fig. 11b).

**Figure 11.**
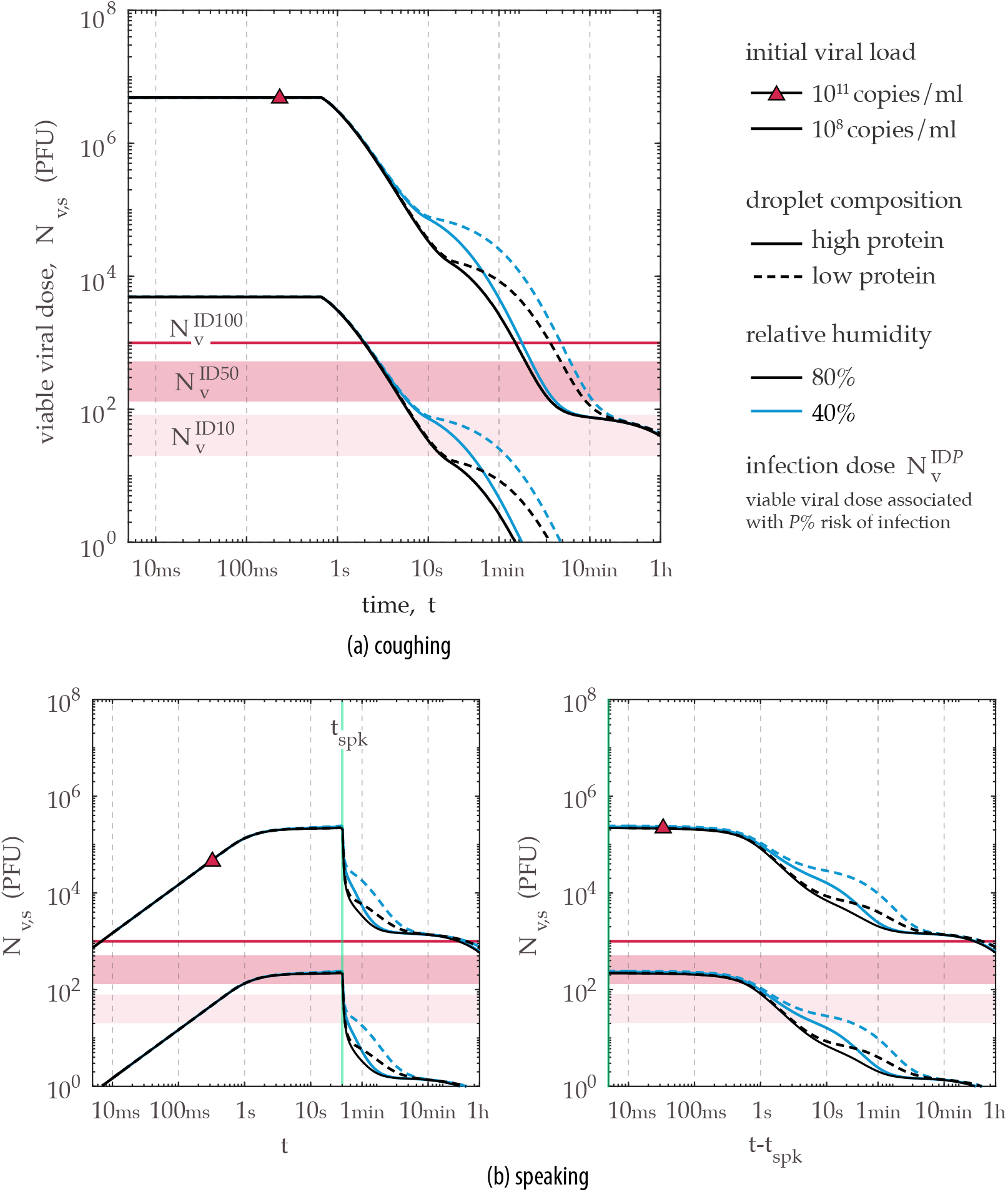
Evolution of the suspended viable viral dose following a short cough in terms of two droplet compositions (low- and high-protein sputum), RH=20 and 80%, and low (10^8^ copies/ml) and high (10^11^ copies/ml) initial viral load *n*_v,0_.

One of the key differences in the decay of suspended virus emitted from (a) coughing in comparison to (b) speaking can be seen in respect to 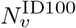 in Fig. 11. For a cough and the worst-case viral load scenario (10^11^ copies/ml_l_), the viral inactivation and removal through gravity will quickly bring the total *N*_s,v_ down to values below those needed for 100% risk of infection (if the whole amount is inhaled by person) in about 1–7 min. In contrast, 30 min are needed for reduction to the same *N*_s,v_ levels in speaking. Due to the small droplets produced in speaking, a high viral dose can still be suspended up to one hour – although this is approximately the half-life of the virus – being one order of magnitude higher than in coughing at that time.

To discuss the evolution of the total number of suspended viable copies of virus in the context of indoors activities, we consider just droplets at face height (between 1.2–1.8 m) in Fig. 12. Based on the results of Fig. 11, here, we assume a worst-case scenario in terms of droplet removal by gravity, that is, a droplet composition of low-protein sputum and relative humidity of 40%. In terms of the virus load in sputum, a typical case of the disease is considered, that is, a high viral load in a patient after a few days of the first symptoms of the disease (*n*_v,0_ =10^9^ copies/ml_l_). Three cases of upward, zero, and downward flow are then evaluated by setting the mean vertical gas velocity as +0.1, 0, and −0.1 m/s. These simple cases aim to illustrate possible flow conditions that may arrive in indoors activities (e.g. thermal plume, heating systems or flow streams caused by ventilation systems). The effect of the mean flow on viral removal at face height is evident: downward flow can reduce the suspended viral dose to safe levels in only a couple of seconds, while upward flow may actually worsen the problem by keeping a higher viral dose suspended at face height for several minutes longer than no flow at all. Moreover, Fig. 12 also illustrates the risk associated with speech-emitted droplets in the short-time scale problem; this will be discussed in detail in the next section, as we consider the gas exhaled during speech.

**Figure 12.**
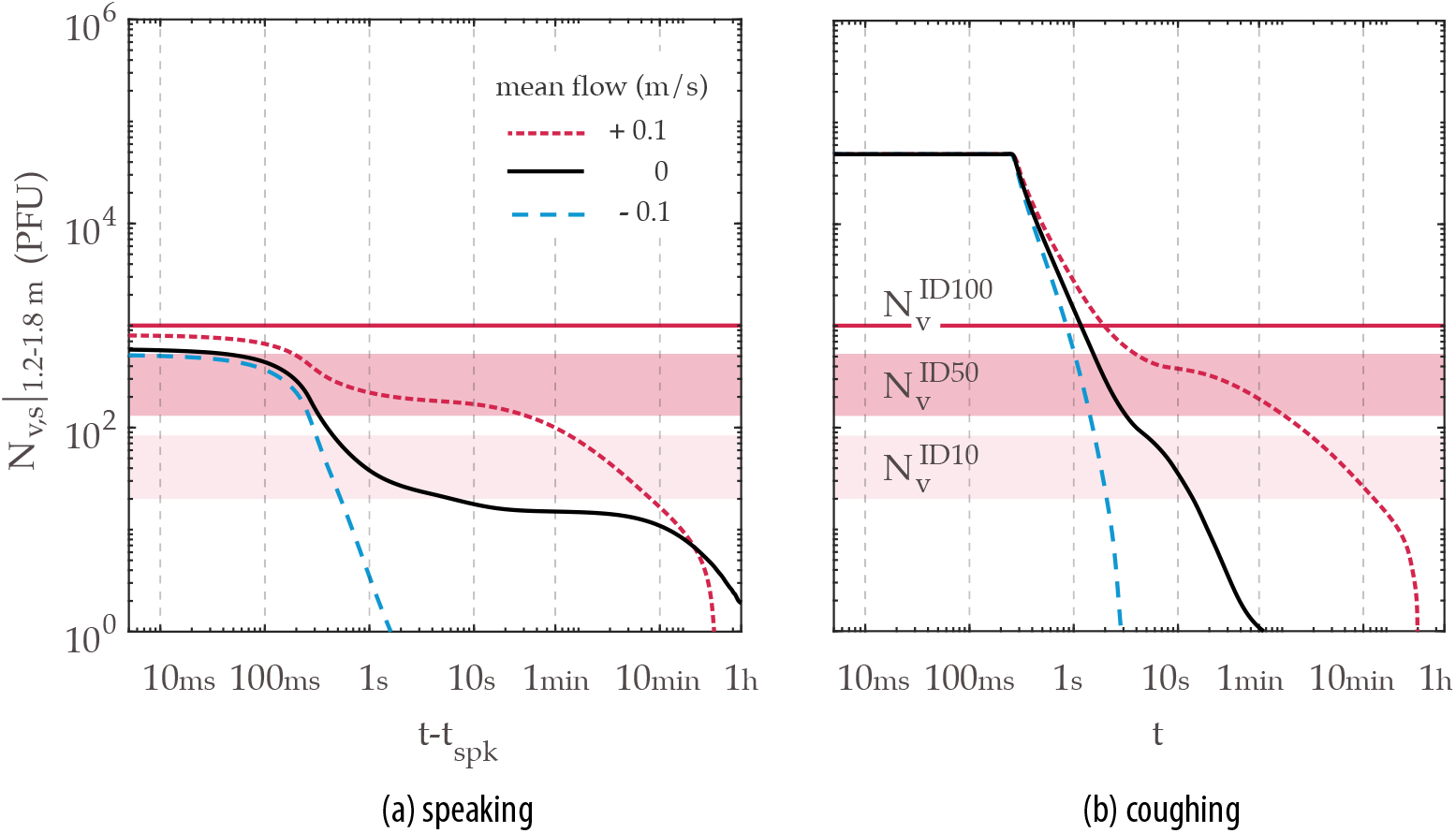
Evolution of the suspended viral dose at face height (between 1.2–1.8 m) following 30-s continuous speech and a short cough (i.e. same exhaled liquid mass) for three cases of ambient mean flow: upward (+0.1 m/s), zero, downward (-0.1 m/s) – low-protein sputum, RH=40%, *n*_v,0_ = 10^9^ copies/ml).

### (d) Implications for physical distancing policies

#### (i) Uniform velocity

In a setting where the initial velocity of the exhaled gas carrying the droplets can be ignored – this might be a good assumption in, say, slow speaking or exhalation in the presence of a strong background airflow – the above time scales can be easily translated into information on space. For example, assuming a uniform velocity *U*_AB_ from emitter A to receptor B located a distance *L* from A in the airflow direction, the total suspended liquid mass can be found by simply setting *t* = *L*/*U*_AB_ in Fig. 6. For instance, for *U*_AB_ = 0.5 m/s and *L* = 2 m, *t* = 4 s and hence about 7 × 10^−3^ to 2× 10^−2^ of the initial liquid is still in the air, depending on the ambient conditions. Similarly, in terms of virus dose, (Fig. 12), an amount up to 100 PFU could be available at face height, depending on specific flow conditions, which illustrates that a 2-m safe distance may not be adequate without the use of protective mask/respirator for the case of strong convection from the emitter to the receptor.

#### (ii) Jet decay

Speaking or coughing involve high velocities at the mouth, which then decay as the flow evolves downstream from the emission source. Here, we model a respiratory emission as a continuous round jet in stagnant ambient air. Although, in reality, the emission is not continuous and the jet is buoyant and behaves like a starting jet of finite duration, the duration of the release, especially for speaking or breathing out, could last of the order of seconds. For such duration, therefore, it may not be a bad approximation to consider the decay of a continuous round jet, which allows us to build an easy-to-use connection between the settling times of Section 3(a) with a time-of-flight estimate to reach the safe distance *L*_sf_. The mean velocity *U* at the centreline of an axisymmetric turbulent jet decays as:

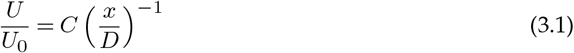

where *L* is the distance from the jet’s source, *D* the jet diameter, *U*_0_ the velocity at the release, and *C* ≈ 6 is an empirical constant from experiment [34]. (The non-circular mouth opening can be absorbed as a first approximation into the value of the constant *C*). Following a fluid particle travelling along the central streamline of the jet,

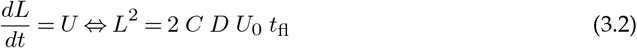

which relates the mean time of flight *t*_fl_ needed to reach a distance *L*. For *L* = *L*_sf_ = 2 m, *U*_0_=5 m/s, *D*=10 mm, values typical of speaking [32], the physical distancing limit *L*_sf_ = 2 m is reached at *t*_fl_ = 6.7 s. The results of Section 3(a) suggest that for speaking more than about 10^−3^ of the emitted release has not yet fallen on the ground at 2 m.

Assume that the steady-state velocity decay (Eq. 3.2) is also valid for a cough. Using *U*_0_=20 m/s and *D* = 20 mm, a distance of *L* = 2 m is reached at about *t*_fl_ = 0.8 s. Figure 11 shows that by that time there has been virtually no gravitational removal when considering the total suspended liquid, and Fig. 12 shows that between 1.2 and 1.8 m, at face height, an order of magnitude removal is observed. Hence, the receptor will receive a significant proportion of all viral load directly at face height. Safe levels are reached only after 30 s–1 min of flight time for either zero or downward flow, which suggests that an unprotected cough is not safe at any reasonable distance. Speaking for 30 s (Fig. 11) with the high-concentration estimate of viral load releases enough virus particles that even after 10 min, the particles that have not settled through gravity are still above the smallest dose needed for 100% risk of infection. However, with the 10^8^ copies/ml_l_ estimate, the suspended viable dose within a few minutes has fallen by about two orders of magnitude. For aerosol droplets in particular, removal by gravity starts only after 20 min with the gravitational settling rate reaching a steady-state value of about 0.39 h^−1^ (0.13 h^−1^ for suspended aerosol droplets by coughing) after 30 min. Both viral inactivation and removal by gravity have a small cleansing effect for the aerosols; hence ventilation removal must be added if continuous speaking is to be safe.

Furthermore, recent work suggests that the voice loudness can enhance droplet production during speaking [35], and that a high viral replication and the highest viral loads of SARS-CoV-2 were found to occur in the throat – where fine droplets are produced – in the early stages of the disease (or in asymptomatic individuals) [4]. Considering such findings in a worst-case scenario of virus loads and potentially greater amounts of droplets emitted in a loud environment, the potential of speaking as a mechanism of indoor as well as outdoor viral transmission becomes even more evident. This corroborates the suggestion that aerosol transmission was the main contagion mechanism in a number of cases, such as the restaurant in Guangzhou (China) and the choir rehearsal in Valley Chorale (USA) as recently suggested in [1].

#### (iii) Mixing ventilation

In the case of mixing ventilation – for instance through ceiling-placed A/C systems, fan-assisted, or jet-induced ventilation – it may be assumed that the air in the room is homogeneously mixed very quickly, typically of the order of the turbulent turnover time *T*_turb_ = *L*_turb_/*u*′, where *L*_turb_ is the integral length scale (perhaps of the order of 0.5–1 m for a typical room) and *u*′the characteristic turbulent velocity (perhaps of the order of 0.02–0.05 m/s). Hence, the mixing timescale may be of the order of 1 to 50 s. This is short compared to a typical purge time of the room by fresh air, which is the inverse of the “air changes per pour” (ACH) that typically characterises ventilation systems. Targets for well-ventilated rooms are 10–20 ACH, which suggests an average purge (or residence) time *T*_res_ of 180–360 s. Therefore, considering the typical ACH used currently, the well-mixed approximation is fair. The *N*_v,s_ would be removed by ventilation according to

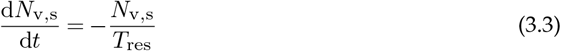

where *T*_res_ = *V*/*Q*, with *V* the volume of the room and *Q* the ventilation volume flow rate. This gives an exponential decay with *T*_res_ as the characteristic timescale.For example, a removal of 10^−4^ of the original *N*_v,s_, associated with the aerosol would be reached at a time *t* ≈ 9 *T*_res_.

Consider an indoor environment with a total volume V=200 m^3^, simulating a lecture room, and one infected individual continuously speaking for 1 h. Let us also assume a range of 10^8^– 10^10^ copies/ml_l_ virus load in sputum. The well-mixed approximation results in an uniform concentration of suspended viral aerosol droplets that are generated by speaking and reduced via simultaneous ventilation removal, viral inactivation and gravity, similar to Fig. 11(b). For an occupant inhaling 0.521 l/s [32], the infection dose model by [31] yields an infection risk of about 0.11–10.7% at the end of emission when the room is ventilated with 1 ACH, whereas the infection risk may increase to 0.18–17% if the occupant spends another hour in the same room. For a well-ventilated room with 10 ACH, the infection risk reduces to 0.03–3.2%, which confirms the importance of ventilation in mitigating risk from long-time emission, especially when the emitter is pre-symptomatic or asymptomatic, i.e. having a lower virus load. However, to ensure a higher cleansing effect and reduce risk to levels well below 0.5% for any virus load in the considered range, one would need O(100) ACH, that is, an order of magnitude stronger than current practice in most residential, professional, and even hospital settings.

## 4 Conclusions

A comprehensive modelling strategy of the evaporation and settling of droplets emitted in respiratory releases (speaking, coughing) was provided in a Lagrangian framework, which included state-of-the-art models for droplet evaporation, effects of the sputum composition on heat and mass transfer, and droplet size distributions. By considering characteristics of the SARS-CoV-2 virus, such as its decay rate, viral load in infected individuals, and estimated infection dose from SARS-CoV-1, theoretical estimates were given for the evolution of the droplet cloud in terms of suspended mass and droplet number and its associated viable viral dose, for a short cough and in continuous, paced speech. Such metrics were then used in the context of canonical problems providing insights on physical distancing and ventilation.

The evolution of the probability density functions of droplet size given by the bronchiolar-laryngeal-oral tri-modal model for speaking and coughing were evaluated. Evaporation time scales of approximately 100 ms and 1 s were observed for droplets within the bronchiolar mode (centred at 1-2 µm) and the laryngeal mode (centred at 6-8 µm), respectively. Large droplets produced in the oral mode (centred at 100-200 µm) were observed to settle by gravity within just a few minutes. Evaporation and settling time scales of the droplets were strongly impacted by the sputum composition, which limited the resulting equilibrium size of the generated droplet nuclei (i.e. “dry droplets”). Varying the droplet composition and the ambient relative humidity, resulted in 90% relative differences in the settling times for droplets up to approximately 100 µm, thus impacting the evolution of the droplet cloud and its associated viral dose.

About one hour after emission, virtually all mass lied in droplets within the aerosol class between 0.4–4 µm in diameter. Although, in absolute terms, a short cough emitted as much liquid mass as 30 seconds of continued, paced speaking, the resulting aerosol contained an order of magnitude more mass for speaking than coughing. Further, the effect of an upward mean ventilation flow was observed to sustain a higher viral dose at face height, increasing to 5 min the time at which an infection dose for 10% infection risk was found suspended, in relation to 1 s and 10 s for speaking and coughing when only gravity was considered. At 10 min after emission, the largest droplets in the cloud were just above 10 µm. These results stress the need for air change through ventilation to reduce the risk of disease transmission in indoors activities, and emphasise the importance of the direction of displacement ventilation and precise account of the aerosol characteristics.

Due to the high associated mass and viral dose in large droplets (∼99% of the emitted value), both a short cough and continued speaking were found to be unsafe within 2 m without personal protective equipment. In this case, the problem is governed by the settling of large droplets (*d*_*k*_ >100 µm) produced in the oral mode, hence the effect of droplet evaporation and composition were negligible. Upward air streams resulting from displacement ventilation could increase the distance travelled by droplets, while downward streams (e.g. from under-floor negative pressure ventilation systems) could be used to enhance droplet removal from face height. As ventilation is of utmost importance for aerosol removal, personal protective equipment is crucial to reduce the risk of short-range contamination in ventilation systems inducing upward flows.

The implications of the present findings for virus transmission control measures are:

- Standing 2 m opposite an infected speaker is not safe without the use of a protective mask or respirator. In the presence of a constant mean flow (*U*_AB_ =0.5 m/s), it was found that up to 100 viable viral copies were within face height after just a few seconds. In the absence of such flow, jet-decay calculations demonstrated that a similar viral dose can be found at face height at the same 2-m distance, corresponding in 10% risk of infection, while for a short cough the suspended dose would lead to 50% risk.
- An unprotected cough is not safe at any reasonable distance close to an infected individual. Jet decay estimates showed that even for an initial viral load of 10^8^ copies/ml_l_, equivalent of that found in infected asymptomatic individuals, safe levels of infection at 2 m from emission are only attained after 30 s–1 min of flight time at face height.
- Settling of the aerosol by gravity was found to be small compared to the viral decay. For the suspended aerosol resulting from speaking, removal by gravity started only after 20 min and stabilised at settling rate of *≈*0.39 h^−1^ after 30 min (0.13 h^−1^ for coughing). Therefore, gravitational settling cannot be relied upon for reducing the infection risk posed by aerosols.
- The presence of air currents strongly affects the suspended viral dose regardless of the exhalation mode. A 0.1-m/s downward flow can remove completely suspended virus in less than 10 s, while a 0.1-m/s upward air stream can maintain at face height a viral charge corresponding to 50% infection risk for *≈* 1 min, and a viral charge corresponding to 10% infection risk for *≈* 10 min.
- An infected person speaking for 1 h in a model room may lead to infection risk levels of 10–20% with inadequate ventilation, but the risk can be reduced by, at least, a factor of three if 10 air changes per hour are employed. However, an order of 100 air changes per hour are needed if the risk for continuous emission is to fall below 0.5% for high viral emission (e.g. from symptomatic emitters). Therefore, ventilation (in terms of both magnitude and direction) is of utmost importance in minimising infection risk indoors.

The present results illustrate the need of addressing the problem between the short and the long time-scale settling, that is, between what is commonly defined as droplet transmission and aerosol transmission (*d*_*k*_ <5 µm, [14]). The concerning time scales observed are 10 s–10 min, in which the droplet cloud is characterised, in addition to the aerosol class, by 5–100 µm droplets. Although these droplets represented just under 1% of the total initial mass and viral dose emitted, this value is roughly 30 times more than what lies in the aerosol class. Within this time scale, effects of composition and of relative humidity played an important role resulting in order-of-magnitude variations of the estimated suspended mass and viral dose.

## Data Availability

All data available within the article is also available upon request.

## Authors’ Contributions

E.M. and P.M.O. conceived the work. All authors contributed to the calculations, analysis, and writing of the manuscript.

## Acknowledgements

The authors thank Dr M.P. Sitte for his assistance with the Abramzon-Sirignano model and also acknowledge Dr Adam Boies (University of Cambridge) for useful discussions.

## Footnotes

1 Mass-transfer equilibrium is achieved in a multi-component evaporating droplet as the vapour pressure exerted by the water at the droplet’s interface is approximately that of the water vapour in ambient air.

2 Airborne transmission is typically defined by the health community as contagion through virus-laden droplet nuclei (i.e. “dry” sputum particles) by a susceptible person [14]. From an aerosol science or fluid mechanics perspective, the term *droplet nucleus* evokes the possibility of hygroscopic growth, that is, water absorption by the particle leading to the formation of a water droplet. This mechanism might, in fact, occur in a sputum-derived droplet nucleus as it undergoes changes in humidity as, for example, while being inhaled [15]. Changes in droplet physico-chemical charactertistics during evaporation, as increase of salts and surfactants [16], and the effects of water reabsorption on the stability of the SARS-CoV-2 virus are not known.

